# Crystallized and Fluid Cognition in Adults Who Stutter

**DOI:** 10.64898/2026.08.19.26360797

**Authors:** Geoffrey A. Coalson, Courtney T. Byrd, Enjoli Richardson, Corrin I. Gillis, Michael Mahometa

**Affiliations:** The Arthur M. Blank Center for Stuttering Education and Research The University of Texas at Austin, Austin, TX; Old Dominion University, Norfolk, VA

**Keywords:** stuttering, intelligence, cognition, stigma

## Abstract

**Purpose:** There is a long-standing perception that individuals who stutter are less intelligent, with the disfluencies unique to stuttered speech often assumed to be the overt reflection of lower intelligence, despite no supporting evidence. The purpose of the present study was to explore the validity of this assumption by examining the cognitive abilities of adults who stutter compared to the general population using the NIH Toolbox^©^ Cognition Battery (NIHTB-CB).

**Method:** Sixty-three adults who stutter completed the NIHTB-CB, which includes seven standardized measures assessing crystallized cognition (Picture Vocabulary, Oral Reading) and fluid cognition (List Sorting Memory Test, Pattern Comparison Processing Speed Test, Flanker Inhibitory Control Test, Dimensional Change Card Sort Test, Picture Sequence Memory Test). The NIHTB-CB generates t-scores adjusted for demographic variables based on a large sample of neurotypical adults.

**Results:** Composite scores of overall cognition for adults who stutter were not statistically equivalent, rather, their scores were higher than the general population. Higher scores were driven by crystallized cognition, with significantly higher scores on Oral Reading subscale.

**Conclusions:** Present findings demonstrate that adults who stutter possess cognitive skills that are comparable to or potentially higher, than the general population. These results challenge the misconception that stuttering reflects diminished intelligence and offer evidence to mitigate stereotype threat.

---

Negative stereotypes toward people who stutter have been documented for decades (e.g., Boyle et al., 2009; St. Louis, 2015; Woods & Williams, 1976). Common perceptions include attributing negative traits to people who stutter, such as being shy, self-conscious, anxious, or nervous (Craig et al., 2003; Dorsey & Guenther, 2000). One particularly persistent stereotype is the belief that individuals who stutter are “less intelligent” (e.g., Amick et al., 2017; Susca & Healy, 2002; Von Tiling & Wolff Von Gudenberg, 2012; Zeigler-Hill et al., 2020). Research investigating cognitive abilities in adults who stutter compared to neurotypical adults suggests that group differences, if detected, are subtle (e.g., < 1 *SD*, see meta-analyses by Ofoe et al., 2018; 2026).

Intelligence is defined as an individual’s global capacity for learning, reasoning, and navigating complex information (e.g., Cattell, 1963; Gottfredson, 1997; American Psychological Association, n.d.). This construct is historically categorized into two components according to Cattell-Horn-Carroll theory of intelligence (McGrew, 2009): crystallized intelligence (i.e., the breadth and depth of an individual’s acquired knowledge and experience) and fluid intelligence (i.e., the capacity for novel problem-solving and adaptive reasoning, supported by cognitive constructs such as executive functioning; Brown, 2016; McGrew, 2009)^1^. Although behavioral and neurophysiological differences in cognitive performance tasks have been inconsistent in individuals who stutter, subtle variations in specific subdomains of cognition – such as phonological working memory – have been observed in prior studies (e.g., Byrd et al., 2012; Byrd et al., 2015). These variations in tasks related to specific aspects of cognition may contribute to differences in within Cattel’s taxonomy of intelligence. Therefore, the purpose of this study examines crystallized and fluid cognitive abilities in adults who stutter using the NIH Toolbox - Cognition Battery (NIHTB-CB; Casaletto et al., 2015; Heaton et al., 2014; Northwestern University and the National Institutes of Health, 2016). The goal is to provide evidence to address the presumed difference between adults who do and do not stutter, with respect to (a) overall cognitive performance, (b) crystallized or fluid cognition, and (c) subdomains of these constructs (i.e., fluid cognition: working memory [list sorting task], processing speed [pattern comparison task], inhibitory control [flanker task], cognitive flexibility [card sorting task], episodic memory [picture sequence task], crystallized cognition: receptive vocabulary [picture naming], literacy [oral reading task]).

## Stuttering stereotype and intelligence

The stereotype linking stuttering with lower intelligence has been reported for decades (e.g., Silverman & Paynter, 1990; Von Tiling & Wolff Von Gudenberg, 2012; Zeigler-Hill et al., 2020). Craig et al. (2003) found that listeners’ opinion of speakers producing 15% stuttering-like disfluencies (SLDs, Yairi & Ambrose, 2005) were described as “low intelligence” and “not very educated” compared to lower stuttering frequencies. Amick et al. (2017) found that listeners rated adults who stutter as having lower cognitive ability based solely on read-speech samples without knowing anything else about the speaker.

Unfortunately, people who stutter may not only be aware of this stereotype, but apply it to themselves (e.g., Boyle, 2013). The pervasiveness of what Craig et al. termed the ‘stuttering stereotype’ is far-reaching, and often internalized, despite the absence of empirical evidence to support this perception.

## Cognition and stuttering

Cattell’s (1963) description of intelligence is comprised of two main subtypes: (1) *fluid intelligenc*e, or one’s innate ability to solve problems, reason, recognize patterns, and navigate abstract thought, and (2) *crystallized intelligence*, or one’s knowledge accumulated via a lifetime of formal or informal education and experiences, such as vocabulary, literacy, culture-specific knowledge, and verbal reasoning (e.g., A is to B as B is to X; see also McGrew, 2009). Most experimental studies of cognition in individuals who stutter have focused on processes underlying fluid cognition, specifically executive functioning, including (but not limited to) working memory, inhibition, cognitive flexibility, and attentional skills (for meta-analysis in children who stutter, see Ofoe et al., 2018; for meta-analysis of adults who stutter, see Ofoe et al., 2026 and Doneva, 2020). Across these meta-analyses, when differences are identified in specific domains for adults who stutter, these differences are subtle, often group differences of less than 1 *SD* (phonological working memory: Hedge’s g = -.57 to -.62; selective attention: Hedge’s g = - .64; dual attention tasks: Hedge’s g = -.30) compared to non-stuttering adults and accompanied by considerable heterogeneity. There are also indications that adults who stutter may exceed non-stuttering adults in certain domains (for variance in tasks measuring working memory and inhibition, see Ofoe et al., 2026, Figures 3, 13).

Examinations of crystallized intelligence in adults who stutter are less frequent and focus primarily on linguistic factors related to oral expression and/or comprehension (e.g., expressive and receptive vocabulary, reading comprehension). Of the available studies, adults who stutter do not demonstrate clear evidence of advanced or depressed vocabulary knowledge (Pellowksi, 2011) nor reading comprehension (Choo et al., 2022; Elsherif et al., 2021). Difficulties have been reported with respect to lexical access under time pressure (e.g., Lescht et al., 2022; Newman & Bernstein Ratner, 2007, cf. Hennessey et al., 2008). Taken together, findings from these child and adult studies indicate any differences, when observed, are subtle variations in fluid or crystallized intelligence.

Moreover, a composite measure of Cattell and Hebb’s description of intelligence – including both crystallized and fluid intelligence – has rarely been examined in adults who stutter.

## Stigma and stigma mitigation

The perception that stuttered speech indicates lower intelligence remains a central component of the stuttering stereotype that is perpetuated in popular media (Evans & Williams, 2015), in political discourse (Hendrickson, 2020; Viser & Arnsdorf, 2024), and by the general public (Bebout & Arthur, 1992; Boyle, 2017; Klassen, 2001) across the lifespan (3–5 years of age: Ezrati-Vinacour et al., 2001; 5–7-year-old: Giolas & Williams, 1958; 9– 11 years of age: Franck et al., 2003; adolescents: Evans et al., 2008; adults: Van Borsel et al., 2011). Although explicit biases are not universal (e.g., Boyle, 2017; Iimura et al., 2018; Werle & Byrd, 2022), the link between stuttering and lower intelligence continues to be a pervasive implicit cultural stereotype. The tendency to view individuals who stutter through this stereotypical lens has been identified by professionals across diverse vocations (e.g., Abu-Dahech & Gabel, 2020; Dorsey & Guenther, 2000; Li et al., 2016; Young et al., 2025), including speech-language pathology (e.g., Anderson & Stuart, 2017; Coalson et al., 2022; Cooper & Cooper, 1996; Walden et al., 2020; Woods & Williams, 1971). This external stigma is often internalized by those who stutter (e.g., Boyle, 2013; Boyle et al., 2023), requiring direct focus during clinical intervention that could potentially be prevented if such stigma did not exist (e.g., Byrd et al., 2024). Because the stigma of stuttering is difficult to dispel both within and beyond the clinic, its veracity warrants direct empirical investigation.

Stigma has long served as a key variable in explaining how societies perceive, maintain, and reproduce stereotypes to uphold social status and hierarchy. Following Goffman’s (1963) seminal work, stigma and stigmatization are defined as processes in which power allows the social branding of body/minds that include elements of labeling, stereotyping, separation, status loss, and discrimination (Darling, 2013; Link & Phelan, 2001, p. 377). This results in individuals who are stigmatized being perceived as undesirable and socially devalued. Critical disability studies further investigate how the social construction of *differences* has been leveraged to maintain the stigma associated with identity. Models of disability and theories seeking to purport racial inferiority related to perceived intellectual and cognitive incapacity have been articulated to justify labeling and discrimination against marginalized groups. Research examining intelligence stigma has been studied across multiple domains, including the racial academic achievement gap (Steele & Aronson, 1995), gender stereotypes (Colom & Garcia-Lopez, 2002; Storage et al., 2020), age stereotypes (Lamont et al., 2021), and disability (Carlson, 2017; Annamma et al., 2013; Daniel, 2025; Fresson et al., 2018; Kumar et al., 2024). Such studies demonstrate how stigma surrounding intelligence operates across social identities and reinforce systemic inequalities through stereotype threat and deficit-based assumptions. Negative reactions as a byproduct of stigma associated with stuttering have contributed to social exclusion and limited opportunities for individuals who stutter due to deficit assumptions surrounding speech communication and competency (Dew & Gabel, 2024; Gabel et al., 2004; Richardson et al., 2025). Amongst stereotypes associated with stuttering, the perception of reduced cognitive intelligence remains uncorroborated and largely unexplored. These outcomes are not isolated but reflective of a broader system of deeply ingrained beliefs about communication, intelligence, and competence. Building on prior studies, the present study extends efforts to confront and mitigate entrenched stigma that equates stuttering with diminished intelligence, thereby reframing stuttering not as a marker of cognitive deficit.

## Purpose of the present study

Decades of research have documented the general public’s perceptions about the intelligence of people who stutter. Although subtle variations in executive functioning in adults who stutter have been found compared to non-stuttering peers, with some finding superior performance in adults who stutter, no study to date has assessed these processes relative to large normative samples of neurotypical adults. If cognitive performance of adults who stutter is found to be comparable to the general population, such data will provide counterevidence to the perceptions about stuttering being the result of lesser intelligence. To examine this, we employed a *statistical equivalence* test (i.e., Two One-Sided Tests, or TOST) to first verify that performance fell within the range of practical equivalence to the general public, with the understanding that if the assumption of equivalence was not met, we would follow up with a traditional Null Hypothesis Significance Test (NHST). Using this sequential statistical approach, the study addressed the following specific research questions:

### Research Question 1 (RQ1)

Does overall cognitive function in adults who stutter differ significantly from the general population?

### Research Question 2 (RQ2)

Do adults who stutter differ significantly from the general population in crystallized or fluid intelligence?

### Research Question 3 (RQ3)

Do adults who stutter differ significantly from the general population on specific subdomains of crystallized or fluid intelligence?

Across RQs, it was predicted that adults who stutter would perform statistically equivalent to the general public, which would refute the pervasive assumption of lower intelligence.

## Methods

### Participants

The study was approved by the authors’ university institutional review board of The University of Texas at Austin (IRB #: 2015-05-0044). Participants provided informed written consent prior to participation. Sixty-three adults who stutter completed the NIH Toolbox – Cognitive Battery (NIHTB-CB; Casaletto et al., 2015; Heaton et al., 2014). The NIHTB-CB test battery was completed in-person either as part of diagnostic assessment and/or previous experimental studies (e.g., Coalson et al., 2025; Gkalitsou & Byrd, 2021).

Stuttering diagnosis was confirmed by a certified, licensed speech-language pathologist with expertise in stuttering at the Arthur M. Blank Center for Stuttering Education and Research at the University of Texas at Austin. Table 1 provides a summary of demographic information across participants.

**Table 1.**
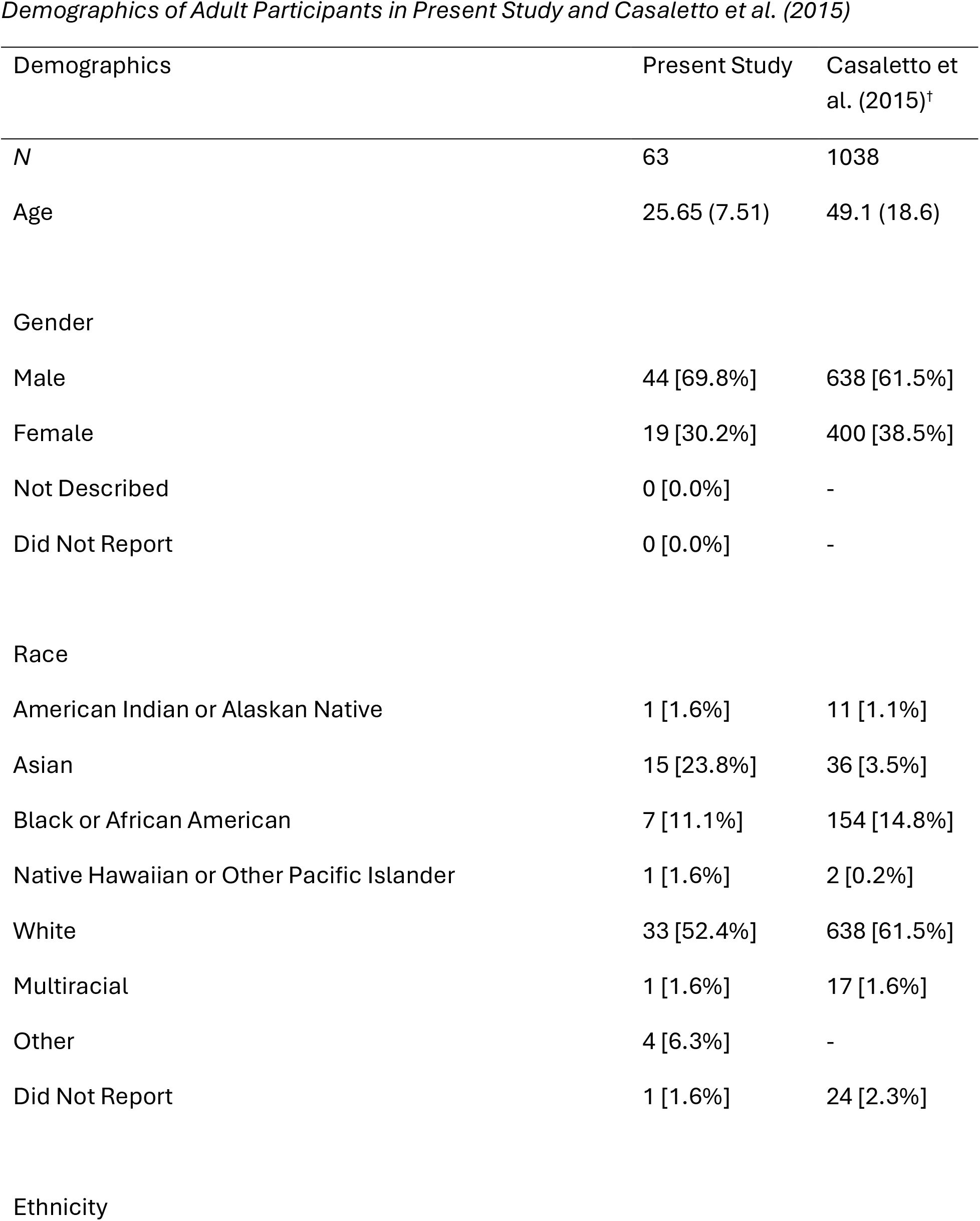
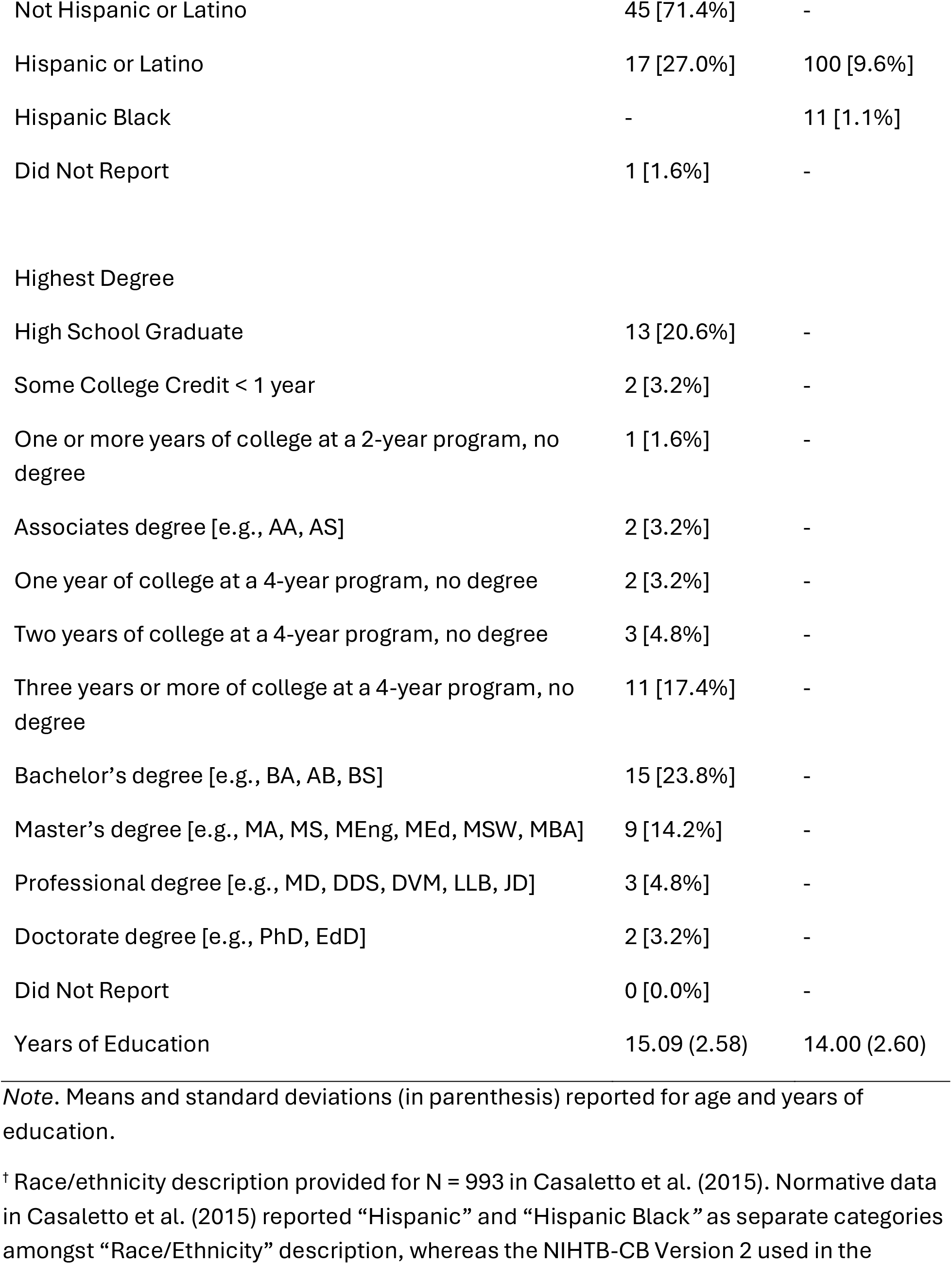
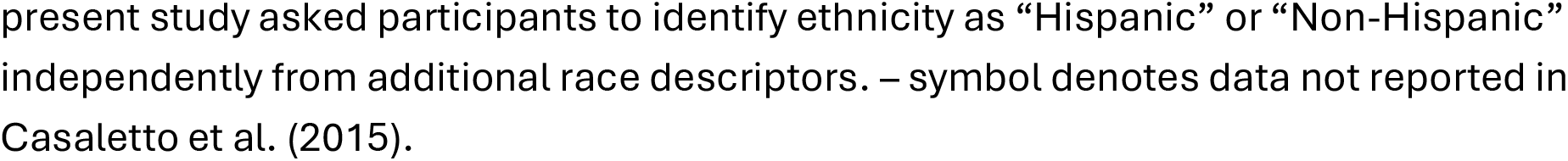
Demographics of Adult Participants in Present Study and Casaletto et al. (2015)

The NIHTB-CB (Version 2; Heaton et al., 2014) is a standardized, iPad-based cognitive assessment consisting of these seven subtests: Picture Vocabulary Test, Flanker Inhibitory Control and Attention Test, List Sorting Memory Test, Dimensional Change Card Sort Test, Pattern Comparison Processing Speed Test, Picture Sequence Memory Test, and Oral Reading Recognition Test (see Table 2 for descriptions of each) that generates a Total Cognition Composite Score, which is derived from both Crystallized and Fluid Cognition Composite Scores and adjusted for five demographic factors: age, education, race, ethnicity, and gender. The Crystallized Cognition Composite Score reflects past learning and accumulated knowledge. The Fluid Cognition Composite Score measures abstract reasoning and problem-solving abilities. Each subtest, as well as the three composite scores, yield a Fully-Corrected T-Score (*M* = 50, *SD* = 10) based on a large national sample (i.e., 1038 neurotypical adults; Casaletto et al., 2015). The Casaletto et al. cohort (described in Table 1) provides the original, core data used to develop the age-, race-, education-, and gender-adjusted T-scores for neurotypical adults for NIHTB-CB Version 2 used by participants in the present study.

**Table 2.**
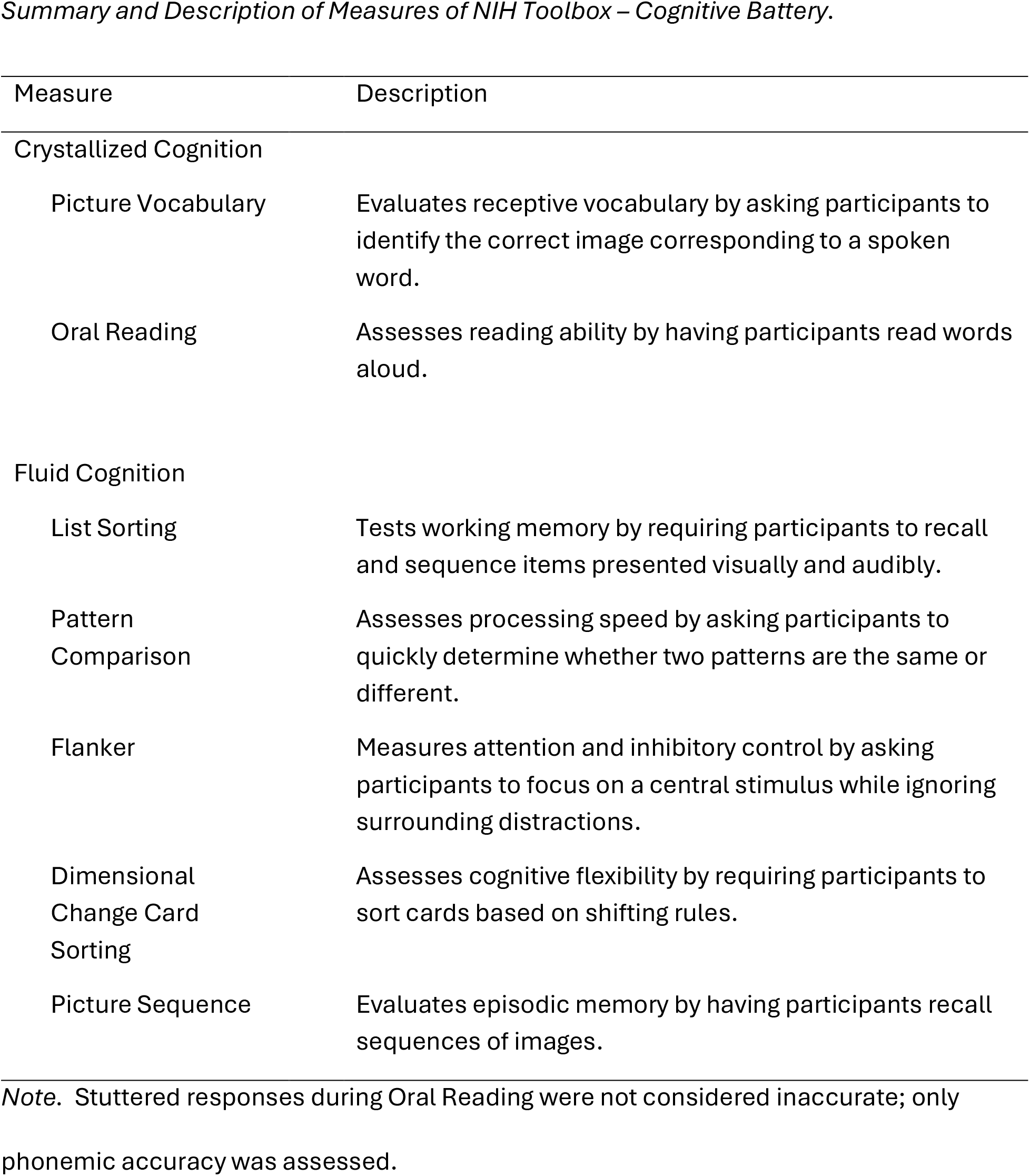
Summary and Description of Measures of NIH Toolbox – Cognitive Battery.

The NIHTB-CB is normed for individuals aged 8 to 85 and takes approximately 30-40 minutes to administer. Participants completed the NIHTB-CB in an isolated room independently, with the exception of two subtests that required examiner interaction (i.e., Oral Reading, Picture Sequence). Examiners for these subtests were research assistants or speech-language pathologists trained in stuttering and familiar with the administration protocol of the NIHTB-CB (Northwestern University and the National Institutes of Health, 2016).

### Analyses

Analyses consisted of two sequential phases across RQ1, RQ2, and RQ3. We first applied the *statistical equivalence* test – the Two One-Sided Tests (TOST) approach – to examine practical equivalence between the sample and the general population. For any comparison where equivalence was not confirmed, we then conducted a traditional one-sided Null Hypothesis Significance Test (NHST).

During both TOST and NHST analyses, we compared data from sample cohort (*N* = 63) to the general population provided by NIH Toolbox normative data (*M* = 50, *SD* = 10, *N* = 1038 adults; Casaletto et al., 2015). Dependent variables across analyses included Fully-Corrected T-Scores of the Total Cognition Composite Score (RQ1), Crystallized Cognition Composite Score and Fluid Cognition Composite Score (RQ2), and scores for each of the seven subtests (i.e., Picture Vocabulary Test, and Oral Reading Recognition Test; List Sorting Memory Test, Pattern Comparison Processing Speed Test, Flanker Inhibitory Control and Attention Test, Dimensional Change Card Sort Test, and Picture Sequence Memory Test; RQ3).

### TOST: Analysis of Statistical Equivalence

In equivalence testing, the null hypothesis is that the difference in means falls outside the equivalence bounds (i.e., the null hypothesis is that the means are different; Lakens et al., 2018). Statistical equivalence for each was determined using TOST approach within the TOSTER package (Caldwell, 2022; Lakens, 2017). Within this approach, scores between samples were considered equivalent when the difference between the sample mean and the population mean fell completely within a confidence interval (i.e., lower bound, upper bound) that is considered sufficiently close to zero to be considered equivalent to zero (see Seidl et al., 2024 for similar reporting and description of TOST statistics).

In order to apply TOST, it was first necessary to determine the upper and lower bounds of the equivalence range around zero. We first calculated the standard effects (Cohen’s *d*) using both population information from NIHTB-CB and sample-based information (*M* = 50, *SD* = 10, *N* = 1038 adults; Casaletto et al., 2015). We then used an approach developed by Hedges and Olkin (1986) to compute a unique 90% confidence interval of the effect size (CI of *d*; Lakens et al., 2018) using effect size calculator for groups of different sizes developed by Lenhard and Lehnard (2022; https://www.psychometrica.de/effect_size.html) for each measure included in RQ1, RQ2, and RQ3. To establish bounds, existing *M*, *SD*, and *N* values from our existing cohort as well as the Casaletto et al. (2015) normative cohort were used as the basis for effect size calculation, resulting in a 90% confidence interval for each scale. The use of a 90% confidence interval is a direct consequence of conducting the equivalence test at the α = .05 level. In the TOST approach, two one-sided tests are performed, each at level α. To reject the null hypothesis of non-equivalence at the 0.05 level, we must demonstrate that the entire (1 - 2α) confidence interval – in this case, 90% – lies within the pre-defined equivalence bounds. Statistical equivalence was therefore considered significant if *p*-values for both the upper bound and lower bound fell below α = .05, or within the 90% confidence interval.

### NHST: Analysis of Significant Difference

If significant equivalence was not detected for a specific measure during TOST analyses, samples were then compared using traditional null hypothesis one-sample *t*-tests, again comparing the sample cohort (*N* = 63) to the general population mean provided by NIH Toolbox normative data (*M* = 50, *SD* = 10, *N* = 1038). Effect size values (Cohen’s *d*; Cohen, 1988) were calculated using *M̅* - *M* / *SD*, wherein *M̅* = sample mean, *M* = population mean of 50, and *SD* = sample standard deviation of 10 (i.e., Casaletto et al., 2015; National Institutes of Health and Northwestern University, 2006-2016). As described by Cohen (1988), *d* of 0.2 = small, 0.5 = medium, 0.8 = large. Prior to both TOST and NHST analyses, data were visually inspected for outliers and, if detected, removed prior to analyses. Bonferroni corrections were applied to accommodate multiple comparisons across subdomains for RQ3.

## Power analysis

A priori power analysis indicated that a minimum sample size of *N* = 34 for each was required to detect medium effect sizes (*d* = .05) with sufficient power (.80, two-tailed, α = .05) when analyzing Total Cognition Composite Score (RQ1), Crystallized and Fluid Cognition Composite Score (RQ2), and individual subtests (RQ3). This calculated sample size for NHST is also valid for the TOST procedure of the equivalence test. The collected sample size exceeds (*N* = 63) this minimum requirement.

## Results

### RQ1: Is overall cognitive function in adults who stutter significantly different from the general population?

Prior to analysis, outliers for Total Cognition Composite scores were identified and excluded if scores exceeded 1.5 interquartile range values (Tukey, 1977; see also Coalson et al. 2024; Werle et al., 2023). Based on Tukey’s threshold for outliers, no participant scores were excluded.

#### TOST

As depicted in Table 3, TOST analysis indicated that adults who stutter were not statistically equivalent to the population mean (lower bound: *p* = .208; upper bound: *p* = .006). That is, adults who stutter scored on average, as a group, 7.56 points higher than the general population (*M* = 57.56, *SD* = 8.65) and exceeded statistical equivalence.

**Table 3.**
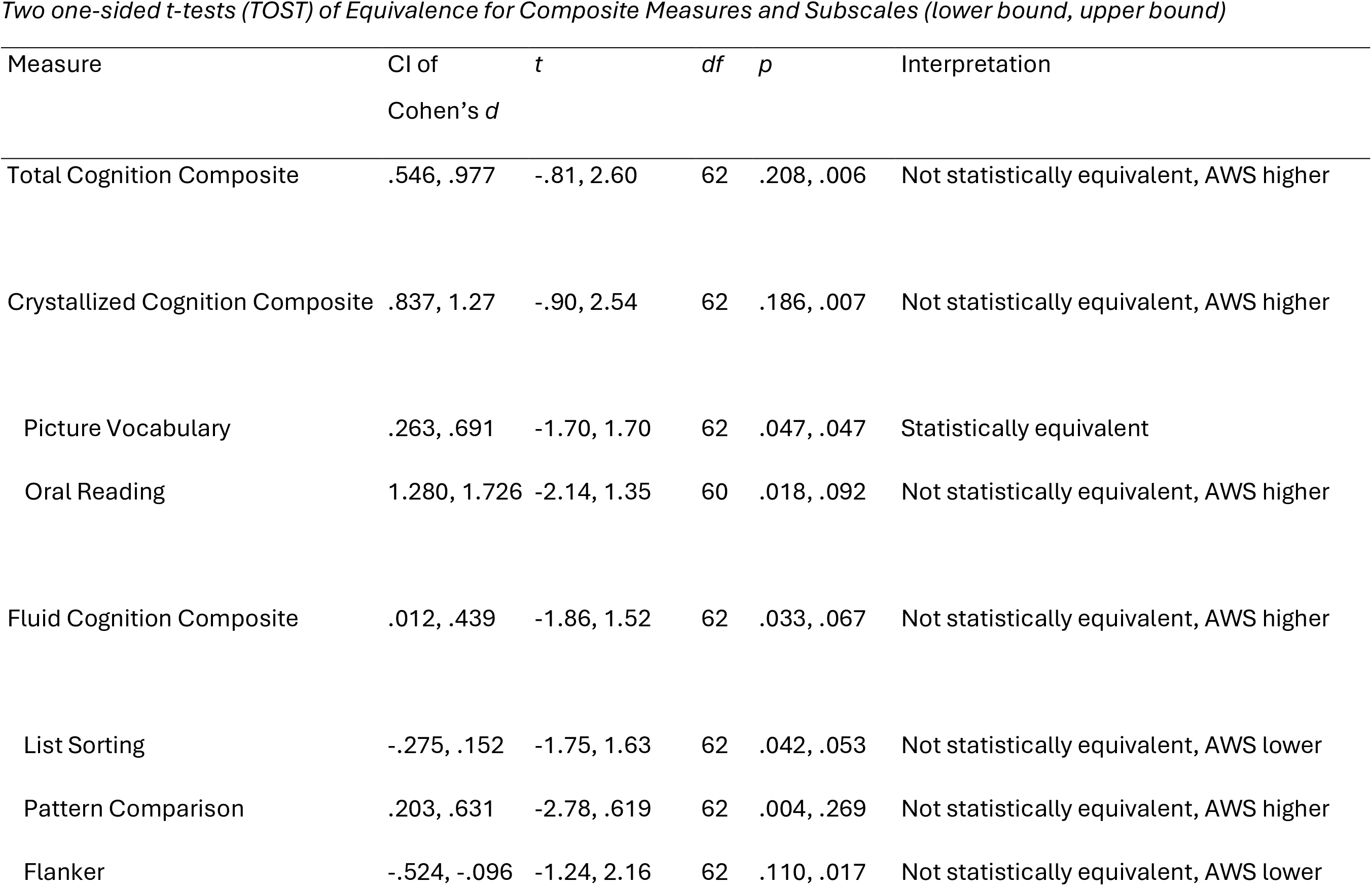
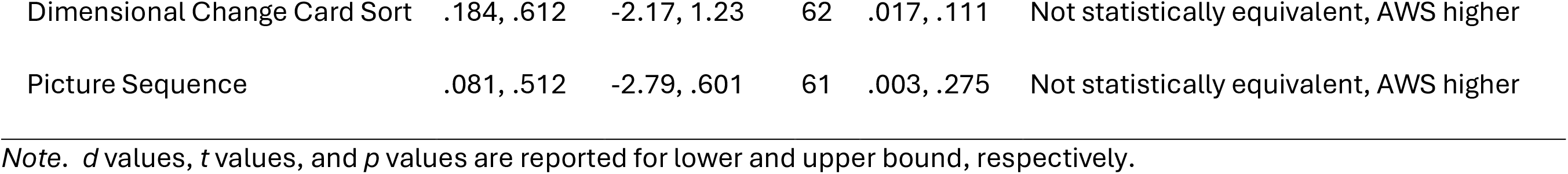
Two one-sided t-tests (TOST) of Equivalence for Composite Measures and Subscales (lower bound, upper bound)

| Measure | CI of<br>Cohen's *d* | *t* | *df* | *p* | Interpretation |
| --- | --- | --- | --- | --- | --- |
| Total Cognition Composite | .546, .977 | -.81, 2.60 | 62 | .208, .006 | Not statistically equivalent, AWS higher |
| Crystallized Cognition Composite | .837, 1.27 | -.90, 2.54 | 62 | .186, .007 | Not statistically equivalent, AWS higher |
| Picture Vocabulary | .263, .691 | -1.70, 1.70 | 62 | .047, .047 | Statistically equivalent |
| Oral Reading | 1.280, 1.726 | -2.14, 1.35 | 60 | .018, .092 | Not statistically equivalent, AWS higher |
| Fluid Cognition Composite | .012, .439 | -1.86, 1.52 | 62 | .033, .067 | Not statistically equivalent, AWS higher |
| List Sorting | -.275, .152 | -1.75, 1.63 | 62 | .042, .053 | Not statistically equivalent, AWS lower |
| Pattern Comparison | .203, .631 | -2.78, .619 | 62 | .004, .269 | Not statistically equivalent, AWS higher |
| Flanker | -.524, -.096 | -1.24, 2.16 | 62 | .110, .017 | Not statistically equivalent, AWS lower |
| Dimensional Change Card Sort | .184, .612 | -2.17, 1.23 | 62 | .017, .111 | Not statistically equivalent, AWS higher |
| Picture Sequence | .081, .512 | -2.79, .601 | 61 | .003, .275 | Not statistically equivalent, AWS higher |

*Note.* $d$ values, $t$ values, and $p$ values are reported for lower and upper bound, respectively.

#### NHST

As depicted in Table 4, NHST analysis one-sample *t*-test indicated that adults who stutter exhibited significantly higher cognitive functioning than the general population, *t*(1, 62) = 6.94, *p* < .0001, *d* = .76 (medium effect size).

**Table 4.** Summary of NIH Toolbox – Cognitive Battery Performance and Statistics.

|  | *M* | *SD* | *Mdn* | *t/Z* | *df* | *p* | *d* |
| --- | --- | --- | --- | --- | --- | --- | --- |
| Total Cognition Composite | 57.56 | 8.65 | 57.95 | 6.94 | 62 | <.001* | .76 (medium) |
| Crystallized Cognition Composite | 60.48 | 9.06 | 62.00 | 9.18 | 62 | <.001* | 1.05 (large) |
| Picture Vocabulary | 54.77 | 10.00 | 54.00 | - | - | - | - |
| Oral Reading | 65.06 | 10.37 | 64.00 | 11.34 | 60 | <.001* | 1.51 (large) |
| Fluid Cognition Composite | 52.27 | 11.15 | 53.00 | 1.62 | 62 | .111 |  |
| List Sorting | 49.39 | 8.84 | 48.00 | -.55 | 62 | .585 |  |
| Pattern Comparison | 54.33 | 15.41 | 55.00 | 2.23 | 62 | .029<sup>†</sup> |  |
| Flanker | 46.85 | 12.49 | 43.00 | -1.99 | 62 | .050 |  |
| Dimensional Change Card Sort | 54.03 | 11.89 | 55.00 | 2.69 | 62 | .009<sup>†</sup> |  |
| Picture Sequence | 53.02 | 12.84 | 51.00 | 1.85 | 61 | .069 |  |

<sup>†</sup>did not exceed Bonferroni-corrected threshold ( $\alpha = .008$ )896 \* $p < .008$ (Holm-Bonferroni-corrected threshold)

### RQ2: Do adults who stutter differ significantly from the general population in crystallized or fluid intelligence?

Similar to RQ1, outliers for Fluid and Crystallized Cognition Composite scores were identified and excluded if scores exceeded 1.5 interquartile range values. No participants exceeded this threshold.

#### TOST

As summarized in Table 3, TOST analysis indicated that adults who stutter were not statistically equivalent to the population mean with respect to Crystallized Cognition Composite Scores (lower bound: *p* = .186; upper bound: *p* = .007). As a group, Crystallized Cognition Scores for adults who stutter were, on average, 10.48 points higher than the general population (*M* = 60.48, *SD* = 9.06) and exceeded statistical equivalence. Similarly, adults who stutter were not statistically equivalent with respect to Fluid Cognition Composite Scores (lower bound: *p* = .033; upper bound: *p* = .067). Fluid Cognition Composite Scores for adults who stutter, as a group, were on average 2.27 points higher than the general population (*M* = 52.27, *SD* = 11.15) and also exceeded statistical equivalence.

#### NHST

As depicted in Table 4, NHST one-sample *t*-test indicated that adults who stutter exhibited significantly higher Crystallized Cognition Composite Scores than the general population, *t*(1, 62) = 9.18, *p* < .001, *d* = 1.05 (large effect size). Adults who stutter do not exhibit significantly higher or lower Fluid Cognition Composite Scores than the general population, *t*(1, 62) = 1.62, *p* = .111.

### RQ3: Do adults who stutter significantly differ from the general population specific cognitive subdomains of fluid and crystallized intelligence?

Similar to RQ1 and RQ2, outliers for each of the six subtests were identified and excluded if scores exceeded 1.5 interquartile range values. Based on this threshold, two participants were excluded from Oral Reading (*N* = 61) and one participant was excluded from Picture Sequence (*N* = 62).

#### TOST

Bonferroni-corrected alpha level of .007 (.05 / 7 subtests) was applied to determine significance. In terms of crystallized cognition, TOST analyses indicated that adults who stutter were statistically equivalent to the population mean with respect to Picture Vocabulary (lower bound: *p* = .047; upper bound: *p* = .047; see Table 3). However, adults who stutter were not statistically equivalent to the population with respect to Oral Reading (lower bound: *p* = .018; upper bound: *p* = .092). As a group, adults who stutter scored on average 15.06 points higher than the general population (*M* = 65.06, *SD* = 10.37), and this difference exceeded statistical equivalence.

In terms of fluid cognition, adults who stutter were not statistically equivalent to the general population across subtests. With respect to List Sorting (lower: *p* = .042; upper: *p* = .053) and Flanker (lower: *p* = .110; upper: *p* = .017), adults who stutter scored, on average, .61 and 3.15 points lower than the general population, respectively, and these differences exceeded statistical equivalence. With respect to Pattern Comparison (lower: *p* = .004; upper: *p* = .269), Dimensional Change Card Sort (lower: *p* = .017; upper: *p* = .111), and Picture Sequence (lower: *p* = .003; upper: *p* = .275), adults who stutter score on average 4.33, 4.03, and 3.02 points higher than the general population, respectively, and these differences exceeded statistical equivalence.

#### NHST

Six of the seven subtests were analyzed using NHST, except for Picture Vocabulary, which was identified as statistically equivalent to the general population. Bonferroni-corrected alpha level of .008 (.05 / 6 subtests) was applied to determine significance. As depicted in Table 4, NHST one-sample *t*-tests indicated that adults who stutter exhibited significantly higher scores than the general population on Oral Reading *t*(1, 62) = 11.34, *p* < .001, *d* = 1.51 (large effect size). Adults who stutter did not exhibit significantly different scores than the general population on the five subtests that comprise the Fluid Cognition Composite Score (*p*-values: .009 to .585).

## Discussion

Stuttering is replete with references in popular media that relate it to diminished intellectual capacity, despite there being no data to support a relationship between the two. Unfortunately, these stereotypes persist and continue to shape public perceptions. The present study sought to compare the cognitive abilities of adults who stutter relative to the general population using the NIH Toolbox Cognition Battery (NIHTB-CB). By examining both fluid and crystallized intelligence, as well as subdomain-level performance, we aimed to clarify whether differences in intellect exist. Preliminary findings suggest that adults who stutter do not exhibit atypically low cognitive processing compared to the general population. In fact, results indicate significantly higher cognitive functioning after adjusting for demographic factors such as age, gender, race, ethnicity, and education. As further discussed below, higher scores were driven by crystallized intelligence.

### RQ1: Overall cognitive function

The first main finding is that adults who stutter scored significantly higher on overall cognition relative to the general population. Results provide empirical evidence to counter the misperception that stuttering is associated with lower cognitive intelligence. From an empirical perspective, it is reasonable to expect comparable cognitive abilities to typically fluent peers, as most behavioral data examining cognitive or executive functioning in adults who stutter indicate no domain-general deficits in cognition. Evidence of higher intelligence, however, was less expected. We note, as discussed below, differences in NIHTB-CB Total scores for adults who stutter were largely driven by domains related to crystallized intelligence (i.e., Oral Reading [*p* <.001]), but further note that higher scores in two of the five domains of fluid intelligence approached significance (i.e., Pattern Comparison [*p* = .029], Dimensional Change Card Sort [*p* = .009]). Combined, these suggest that the overall cognitive profiles of adults who stutter may reflect strengths in multiple underlying constructs of executive functioning relative to the general population.

Although outcomes from the NIHTB-CB are encouraging, these outcomes should be interpreted with caution with respect to standardized measures of intelligence (i.e., *Wechsler Adult Intelligence Scale - 4th Edition* [Wechsler, 2009], *Stanford-Binet Intelligence Scale* [Roid, 2003], *Woodcock-Johnson – 4^th^ Edition* [Schrank et al., 2018]). The NIHTB-CB shares considerable variance with these traditional tests of generalized intelligence, but these measures are not redundant. In general, NIHTB-CB Total

Composite Score is highly correlated with such ‘gold standard’ measures (*r* = .78; Heaton et al., 2014 [*n* = 268]; *r* = .89; Hessl et al., 2016 [*n* = 63]). Heaton et al. (2014) found that, on average, scores on NIHTB-CB were nearly identical to criterion standard measures for adults who completed both (*n* = 89; *M* = 10.0, *SD* = 2.9 and *M* = 9.9, *SD* = 2.9, respectively). However, while high performance on the specific NIHTB-CB domains, as observed in the present study, is necessary for high intellectual functioning, it is not sufficient to guarantee equivalent scores on generalized intelligence measures. It is possible, for example, that an individual performs strongly on the NIHTB-CB but underperforms on generalized measures of intelligence. Such discrepancies, in theory, would indicate participants’ difficulties in higher level cognitive tasks such as verbal abstraction and multi-step reasoning (as well as potential differences related to test format or measure-specific linguistic and cultural biases). Although generalized intelligence scores were not available for this sample, performance on the NIHTB-CB nevertheless provides evidence of similarities as well as distinct strengths in adults who stutter, relative to the general population, across cognitive domains essential to support general intellectual functioning. These findings challenge deficit-based assumptions with evidence that is needed to mitigate the stigma surrounding this population’s cognitive capacity.

### RQ2: Crystallized versus fluid intelligence

The second main finding is that adults who stutter scored significantly higher in crystallized intelligence than the general population and comparable to the general population in fluid intelligence. Given that crystallized intelligence is thought to reflect knowledge acquired from the environment, typically by virtue of formal education, a straightforward interpretation of this outcome implies that the sample of adults within this study received greater and/or higher quality education. Further, although the NIHTB-CB Fully-Corrected T-Score are adjusted for participants’ years of formal education, the high proportion of advanced college degrees within this cohort (27/63 participants, or 43%) is notable and, in theory, may exceed the adjustment range of the normative model.

However, comparison of the present cohort to the Casaletto et al. (2015) normative sample reveals that the mean years of education in the study sample (15 years) is remarkably similar to the normative mean of the NIH cohort (14 years; see Table 1). Comparable educational attainment between the sample and the normative group, in addition to use of the Fully-Corrected age-, education-, race/ethnicity, and gender-adjusted T-scores, further mitigates concerns regarding educational bias. Findings provide greater confidence that the performance observed in adults who stutter reflects authentic cognitive abilities rather than factors related to unequal education.

Fluid intelligence, on the other hand, is largely resistant to environmental or educational influence, and more sensitive to neurobiological factors such as aging, neurological injury, and clinical factors such as depression and anxiety. The fact that fluid intelligence was descriptively higher than the general population (*M* = 52.67), although not statistically higher (*p* = .111), suggests that fluid cognition is (a) not atypically low in adults who stutter, and (b) these above-average scores are not reflective of environmental or experiential variations. Taken together, our findings suggest, at minimum, that adults who stutter exceed statistical equivalence in more latent aspects of cognition (fluid intelligence), and may potentially excel in dynamic domains of intelligence (crystallized intelligence).

The disparity between fluid and crystallized scores is not without precedent. Certain cultural populations demonstrate elevated crystallized intelligence alongside average or attenuated fluid reasoning - a dissociation that may reflect the differential impact of life experiences on specific cognitive abilities of this specific sample of adults and underscores the importance of interpreting cognitive profiles within sociocultural context. As observed in the present study, certain populations demonstrate elevated crystallized intelligence alongside average fluid reasoning (e.g., school-age children with greater intellectual engagement [e.g., Ganzach, 2021; Hülür et al., 2018; cf. Bergold & Steinmayr, 2024]; older adults with stronger cognitive reserves [e.g., Tucker-Drob et al., 2009; cf. Tucker-Drob et al., 2022]). Crystallized intelligence is shaped not only by formal education, but by factors which can vary independently from formal education, such as cultural exposure, reading habits, academic engagement, and verbal enrichment.

Although these were not measured in the present study, future research should further explore the potential contribution of informal education and experiences to differences in cognitive strengths across the groups.

To account for age-related variation, all Fully-Corrected T-Scores provided by NIHTB-CB were adjusted against age-matched norms. It is notable, then, that despite our cohort (*M* age = 25) being younger than the Casaletto et al. (2015) sample (*M* age = 49; see Table 1), they still performed above the normative mean in crystallized cognition. To ensure these age-adjustments were not artificially inflating or masking results, we reviewed Uncorrected Standard Scores. These uncorrected scores also indicated an average score of 110.54 (*SD* = 6.77) were functionally comparable to test norms (*M* = 100, *SD* = 15). These findings counter deficit-based models, suggesting instead that crystallized cognition in adults who stutter are comparable to the general public.

### RQ3: Subdomains of crystallized or fluid intelligence

#### Crystallized intelligence

Our third main finding was that adults who stutter scored significantly higher than the general population on one measure of crystallized intelligence. In specific, for Oral Reading, AWS were significantly stronger than the general public (see Table 3). Oral Reading requires print-to-sound transformation and verbal output, making it a literacy- and decoding-sensitive indicator of crystallized cognition. This finding of Oral Reading being stronger for AWS may be surprising given the subtle difficulties in phonological encoding documented in adults who stutter compared to neurotypical peers (e.g., Bosshardt, 1990; Byrd et al., 2015; Sasisekaran et al., 2006; Sasisekaran et al., 2013; Weber-Fox et al., 2004).

On the NIHTB-CB, the Oral Reading Recognition Test primarily reflects reading decoding and print-based expressive language: it requires the integration of orthographic processing (recognizing letter and word forms), phonological decoding (mapping print to sound), and articulation of words, including irregular and low-frequency items. In this sense, high Oral Reading performance suggests efficient use of learned grapheme– phoneme correspondences, automatized word recognition, and literacy-linked crystallized abilities, which are often shaped by educational history and reading experience. Thus, Oral Reading reflects a literacy- and decoding-sensitive indicator of crystallized cognition.

Future research should aim to expand the educational profile of participants to determine to what degree education may contribute to the outcomes, as well as recruit a propensity sample of non-stuttering adults matched by education.

#### Fluid intelligence

With respect to fluid intelligence, although we did not predict significant differences, it would be reasonable to have found lower scores on List Sorting subscale given the documented subtle, < 1 *SD* differences in phonological working memory (e.g., Byrd et al., 2012; Byrd et al., 2015; Coalson & Byrd, 2018; Sasisekaran, 2013; Sasisekaran & Weisberg, 2014). On the contrary, findings indicate that adults who stutter scored similarly to the general public on the task of working memory (i.e., List Sorting Working Memory subtest; *M* = 49.39, *SD* = 8.84, *p* = .585). That being said, the List Sorting subtest is a domain-general working memory task requiring the participant to sequence lexical, semantic items by category and size (e.g., apple, elephant, strawberry, dog…). As noted by Ofoe et al. (2026), adults who stutter typically exhibit difficulties retaining non-lexical stimuli during tasks such as nonword repetition. Thus, the List Sorting task may not have isolated phonological processing in a manner that would expose underlying difficulties in phonological working memory. Moreover, differences between fluid intelligence for adults who stutter were characterized not by lower scores on working memory subtest, but by stronger scores on subtests not directly related to working memory (i.e., processing speed: Pattern Comparison; cognitive flexibility: Dimensional Change Card Sort; episodic memory: Picture Sequence).

Finally, it is noteworthy that several measures within fluid intelligence were higher than the general population mean, each nearing a half a standard deviation above the mean T-Scores (i.e., 55) and either approached or exceeded the Bonferroni-corrected threshold (α = .008; Pattern Comparison [*M* = 54.33, *SD* = 15.41, *p* = .029], Dimensional Change Card Sort [*M* = 54.03, *SD* = 11.89, *p* = .009]). Although we applied a conservative estimate of significance, the findings suggest that increased sample size and statistical power may reveal underlying strengths in fluid intelligence unrelated to the lived experiences of stuttering and/or formal education (i.e., processing speed, cognitive flexibility, and episodic memory, respectively). Only one subtest of fluid intelligence - the Flanker task – approached significance with a score below the general public (*M* = 46.85, *SD* = 12.49, *p* = .050). Further investigations are justified to assess such differences, although similar to working memory differences, it is likely that inhibition differences in adults who stutter, if detected, are subtle (e.g., <1 SD).

## Clinical Implications

The importance of dispelling pervasive myths about stigmatized populations cannot be understated (e.g., McKown & Weinstein, 2003; McKown & Stambler, 2009). Too often, the negative impact of daily implicit (and oftentimes explicit) messages regarding stuttering as a mental deficit is overlooked as an actionable clinical goal. Taking the time to provide the child or adult who stutters, and their families, with accurate information to challenge such beliefs is an integral part of stuttering-affirming, strengths-based models of treatment (e.g., *Education* subcomponent of CARE Model approach; Byrd et al., 2024).

Findings from the present study provide clear evidence for clinicians to say to a client, explicitly: ‘Research shows that when we test the intelligence of people who stutter, they do just as well, and in some cases better, than people who do not stutter’.

Acknowledgement of this by the clinician, caregivers, and the person who stutters helps to prevent emerging internalized stigma or dismantle existing negative beliefs.

## Considerations and Future Research

Several considerations should be noted. First, although sufficient statistical power was obtained to provide meaningful outcomes, additional participants with demographics more proportional to the general population would benefit future studies and inform the potential influence of education. Second, the normative adjustment procedures of Fully-Corrected T-Score calculated by the NIHTB-CB warrant consideration. Our cohort of participants completed NIHTB-CB Version 2, which adjusts the Fully-Corrected T-Score based on the participant’s race, gender, ethnicity, age, and education. The newer version of the NIHTB-CB (Version 3, released in 2024; Hook & Giella, 2024; LaForte et al., 2024) omits adjustments based on race and gender – a change these authors endorse.

Unfortunately, it was not possible to isolate age- and education-adjusted scores from race- and gender-adjusted scores in Fully-Corrected T-Scores of Version 2. The choice to use the Fully-Corrected T-Score in the present study, although not ideal, was to adjust, to the extent possible, for age and education which, unlike gender and race, remain relevant, particularly given the present study’s outcomes. However, future researchers are encouraged to use measures such as NIHTB-CB Version 3 to minimize unnecessary demographic adjustments and enhance the precision of cognitive assessments.

## Conclusion

Taken together, the present findings provide evidence that adults who stutter demonstrate cognitive performance that is not inferior to that of the general population. These results offer a data-driven counterpoint to stereotypes equating stuttering with reduced intelligence. The observed strengths in crystallized intelligence may be shaped by educational attainment and/or the complex interplay between lived experience and cognitive development. Future research should continue to explore these associations with the aim of mitigating stigmatization through advancing understanding of the cognitive capacities of adults who stutter.

## Data Availability

All data produced in the present study are available upon reasonable request to the authors.

## Acknowledgements

The authors thank Katelyn Alcott, Sarah Bunyan, Jack Donahoo, Hudson Hanna, Zaniel Florez, Valarie Morales, Grace Morawiec, and Diana Perez for their tireless efforts in data collection and data management. Finally, this study would not have been possible without each person who stutters who shared their time. Thank you for sharing your voices and stories, and for helping to advance assessment, treatment, and quality of life for people who stutter worldwide.

## Data Availability Statement

Datasets are available upon request.

## Footnotes

1 The Cattell-Horn-Carroll theory organizes intelligence into hierarchical strata, from broad to narrow: (1) general intelligence (or *g*), (2) fluid intelligence (*Gf*) and crystallized intelligence (*Gc*), and (3) narrow cognitive abilities (McGrew, 2009). The NIH Toolbox Cognition Battery (NIHTB-CB) provides composite scores for fluid cognition and crystallized cognition derived from measurable behaviors on specific tests known to reflect Gf or Gc, respectively (Akshoomoff et al., 2013). Thus, in the present study, the terms crystallized *cognition* and fluid *cognition* will be used in reference to performance on the NIH Toolbox, with understanding that performance on these tasks reflect broader constructs of crystallized *intelligence* (Gc) and fluid *intelligence* (Gf), which combined are a proxy for a general factor of *intelligence* (g).

